# Sensitivity Analysis and Dynamical Behavior of an Atangana-Baleanu-Caputo Fractional SEIRV Model: A Case Study of the 2004-2005 H3N2 Influenza Season

**DOI:** 10.64898/2026.01.26.26344824

**Authors:** Taylan Demir, Hasan Hüseyin Tosunoğlu

## Abstract

This study presents a theoretical and mathematical framework for understanding the dynamical behavior of infectious disease spread using a compartmental modeling approach. The proposed model incorporates memory effects to capture temporal dependencies that are not adequately represented by classical formulations. Qualitative analysis is employed to investigate the stability properties of the system and the role of key mechanisms in shaping long term dynamics. Publicly available surveillance information is used only to illustrate the consistency of the model behavior with observed trends. The results highlight the value of memory based modeling structures for describing complex biological processes and provide a general mathematical perspective for studying epidemic dynamics.

## 1 Introduction

Seasonal influenza A (H3N2) is a serious health concern for public health agencies and world populations and causes a significant degree of illness and death [1]. H3N2 has the highest degree of antigenic drift among the influenza virus subtypes and can escape vaccine-induced immunity [2]. The 2004–2005 influenza season represents an example in which H3N2 dominated the U.S. influenza activity; laboratory confirmed cases of H3N2 accounted for a major proportion of the cases reported in this season [3]. The large number of influenza-like illnesses (ILI) reported in the months between November 2004 and March 2005, as shown by CDC data, created an opportunity to utilize mathematical methods to study the dynamics of H3N2 spreading and its dynamics. H3N2 influenza A is a strain of seasonal flu that is a significant global public health problem that results in increased rates of hospitalization and death [1]. Unlike other strains of influenza A, H3N2 experiences increased levels of antigenic drift that can lead to immunity escape, resulting in lower vaccine efficacy compared to other strains of influenza A [2]. The 2004-2005 influenza season was marked as the year that H3N2 was named the dominant strain of the flu virus in the USA, with over half of the laboratory-confirmed cases attributed to H3N2 [3]. According to CDC data, the substantial increase in the number of Influenza-Like Illness (ILI) related visits visiting the healthcare system during the peak of the influenza season is also an important source of mathematical modelling related to the transmission dynamics of ILI during the influenza season.

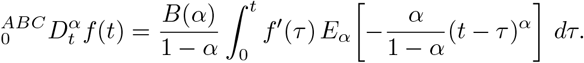

The normalization function is represented by *B*(*α*) and the Mittag-Leffler function is denoted by *E*_*α*_ [5]. Although the traditional SEIR structure comprises of four compartments: Susceptible (*S*), Exposed (*E*), Infectious (*I*), and Recovered (*R*), there is also a need to have a vaccinated compartment (*V*) when analyzing viruses, such as *H*3*N* 2, where vaccination will play an important part in controlling the spread of the disease [6]. Consequently, a fifth compartment in the form of SEIRV allows for a clear assessment of vaccine-induced immunity in generating breaks in chains of transmission as well as the resultant effects on *R*_0_ the basic reproductive number. This investigation into the behavior of an ABC–SEIRV model based upon *H*3*N* 2 data from the 2004–2005 season will explore, quantitatively through means of sensitivity analysis, the most efficient strategies for controlling disease outbreaks.

## 2 Preliminaries

The mathematical definitions of the Atangana-Baleanu-Caputo (ABC) operator used to model the H3N2 virus’s transmission behaviour are provided in this section [5]. The ABC fractional derivative operator uses a memory effect to determine a system’s past states (e.g. previous infections) in contrast to the classical Caputo derivative [4,5] that does not account for past states in its calculations. By using an Mittag–Leffler kernel [7], the ABC fractional derivative operator contains both non-local and non-singular properties [5], giving the expression a high degree of fidelity to sudden changes and complex behaviours that occur within biological systems as a result of their many changing variables and interactions.

### Definition 1

Let *f* ∈ *H*^1^(*a, b*) and *α* ∈ [0, 1]. The fractional derivative in the Atangana-Baleanu-Caputo sense is defined by [5];

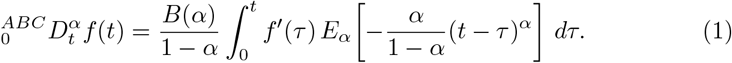

Here, *B*(*α*) is a normalization function satisfying *B*(0) = *B*(1) = 1.

### Definition 2

The one-parameter Mittag-Leffler function appearing in (1), which characterizes the damping behavior of the system is given by its series expansion [7]:

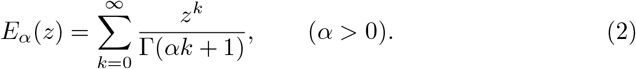

### Definition 3

For a function *f* (*t*), the Atangana–Baleanu (AB) fractional integral introduced as the inverse operator of the ABC derivative is defined by [5];

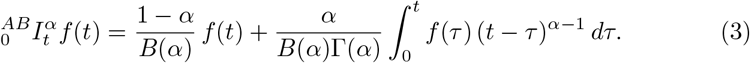

The fractional order *α* reflects the model’s “memory depth,” and as *α* → 1 the formulation approaches the classical integer-order differential system. This flexibility makes it possible to reproduce the 2004–2005 season H3N2 case trends reported by the CDC with a closer match to observed dynamics [3].

## 3 SEIRV Model Formulation

A mathematical representation of the spread of *H*3*N* 2 during 2004 and 2005 was formulated based on data that had been collected at that time, including those obtained from the Centers for disease control [1,2,3]. The mathematical model divides the entire population into five different categories [6] to illustrate how they interact with one another as well as their response to illness and how illnesses are spread.

### 3.1 Definition of Variables

The population classes used in the model are defined as follows [6]:

*S*(*t*) : represents susceptible (unvaccinated) individuals.

*E*(*t*) : denotes individuals exposed to the virus but not yet infectious.

*I*(*t*) : represents infected and infectious individuals; this class corresponds to the cases shown by the red curve in the shared plots.

*R*(*t*) : includes individuals who recover from infection and acquire immunity.

*V* (*t*) : denotes the vaccinated population that is protected against the virus.

### 3.2 ABC–SEIRV System of Equations

Using the Atangana–Baleanu–Caputo (ABC) fractional operator, the governing system of equations is given by [5,6]:

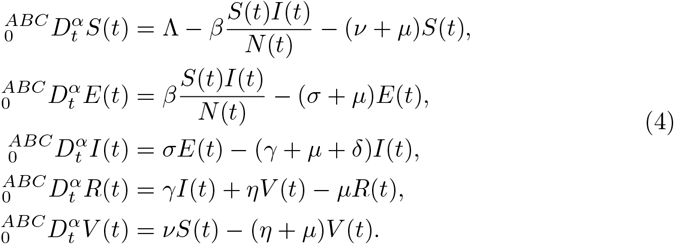

### 3.3. Parameter Analysis and Attribution

The flow dynamics of the model are governed by the following parameters [3,6,7,8]:

- Λ: the rate at which new individuals enter the system (birth/immigration).
- *β*: the transmission coefficient of the H3N2 virus.
- *ν*: the vaccination rate of the susceptible population; this parameter is directly tied to vaccination strategies during the 2004–2005 season.
- *σ*: the progression rate from the exposed (latent) stage to the infectious class *I*.
- *γ*: the recovery rate of infected individuals.
- *µ*: the natural mortality rate in the population.
- *δ*: the influenza-induced mortality rate (disease-related deaths).
- *η*: the rate at which vaccine protection wanes over time, or equivalently, the transition rate of vaccinated individuals into the recovered class.

## 4 Stability Analysis

In this section, we analyze the mathematical consistency of the ABC–SEIRV model [5] and examine the epidemic threshold conditions [9, 10].

### 4.1 Existence and Uniqueness

To establish the existence of solutions, we transform the system into an equivalent integral form and apply fixed-point theory. The SEIRV model under consideration can be written in the operator form [5]

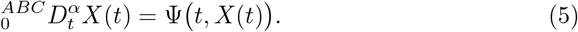

Here, *X*(*t*) = (*S, E, I, R, V*)^*T*^ denotes the state vector. Applying the AB fractional integral to (5) yields the following integral equation [5]:

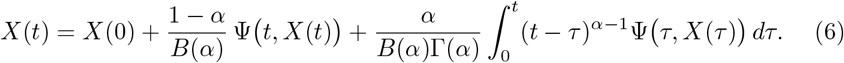

#### Lipschitz Condition

To show that the mapping Ψ satisfies a Lipschitz condition, we consider two solutions *X* and *X*^∗^. Since the components of Ψ (for instance, terms such as *βSI/N*) are differentiable on a bounded region, there exists a constant *L* > 0 such that

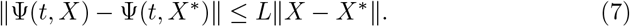

#### Picard Operator

We then define a Picard operator *P* and verify that it is a contraction. If the condition

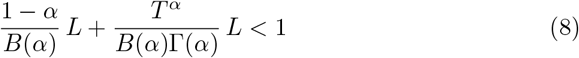

holds, the Banach Fixed-Point Theorem guarantees that the system admits a **unique** solution on the interval [0, *T*].

### 4.2 Computation of the Basic Reproduction Number (*R*_0_)

The basic reproduction number *R*_0_, which quantifies the transmission potential of the H3N2 virus in the population, is derived via the next-generation matrix approach by focusing on the infected compartments (*E* and *I*) [9,10].

**Step 1:** Construction of the *F* and *V* matrices. The matrices *F* and *V* are formed by considering, respectively, the rate at which new infections enter the system and the transitions between compartments (e.g., recovery and death) [9,10]. They are given by

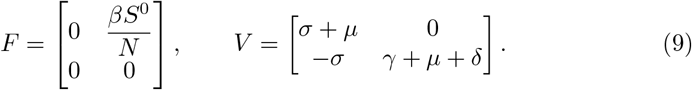

Here, *S*^0^ denotes the susceptible population at the disease-free equilibrium (DFE).

**Step 2:** Computation of the spectral radius. The basic reproduction number *R*_0_ is given by the dominant eigenvalue of the next-generation matrix *K* = *FV* ^−1^ [9,10]. In particular, we have

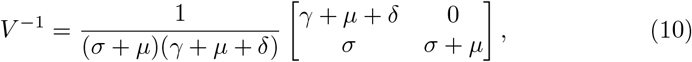

and hence [9,10]

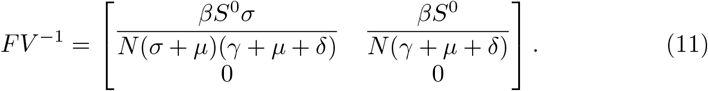

Taking the spectral radius *ρ*(·) of this matrix yields the explicit formula [9,10]

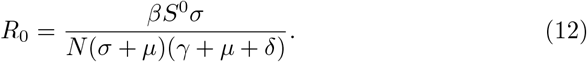

### 4.3 Stability Analysis

To understand the model’s long-term behavior, we examine the stability of both the disease-free equilibrium (DFE) and the endemic equilibrium (EE) [6,10].

#### 4.3.1 Local Stability of the Disease-Free Equilibrium (DFE)

The disease-free equilibrium of the system is defined as

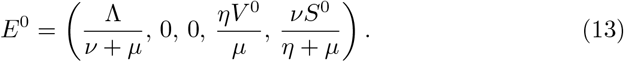

To determine the local stability of this equilibrium, we consider the Jacobian matrix *J* of the system. For a fractional-order system to be locally asymptotically stable, all eigenvalues *λ*_*i*_ of the Jacobian must satisfy the [8,12]

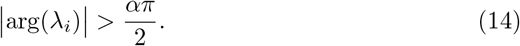

When the ABC operator is employed, the location of eigenvalues in the complex plane typically yields a broader stability region than in classical integer-order models [5,12]. In particular, if *R*_0_ < 1, then all roots of the characteristic equation of the Jacobian meet the above criterion, implying that the epidemic dies out in the population [9,10].

#### 4.3.2 Global Stability Analysis

To prove that the system converges to the DFE regardless of the initial conditions, we construct a Lyapunov function *L* [11]. For the H3N2 model, a suitable Lyapunov candidate is chosen as [11]

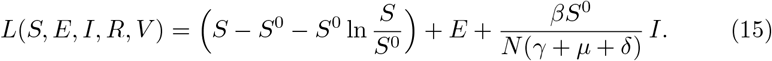

Taking the ABC fractional derivative of this function and imposing the condition *R*_0_ ≤ 1, we obtain [5,11]

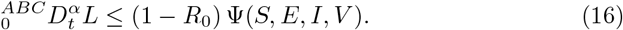

Here, Ψ is a positive definite function. When *R*_0_ ≤ 1, it follows that 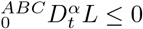.By LaSalle’s invariance principle, the DFE is globally asymptotically stable [11]; that is, if vaccination and control measures are sufficiently effective (*R*_0_ ≤ 1), the outbreak is brought under control throughout the population, independent of initial conditions [10,11].

### 4.4 Relationship Between the Fractional Order *α* and Stability

The ABC operator parameter *α* affects how fast stability is reached [5]. The Mittag-Leffler decay means smaller values of *α* will show a power-law behaviour towards equilibrium rather than the classical exponential behaviour usually expected [5,7,12]. This is why the decline in the number of cases seen in the *H*3*N* 2 case data from 2004 − 2005 differs from what classical exponential models predict [12].

## 5 Sensitivity Analysis

Normalized sensitivity indices were used to find out which factors impact the basic reproduction number *R*_0_ the most [10], using 2004-2005 seasonal data [3] to mathematically justify the prioritization of specific public health interventions [3,6,10].

### 5.1 Definition of the Normalized Sensitivity Index

For the threshold quantity *R*_0_, the sensitivity index with respect to any parameter *p* measures the proportional impact of a small change in *p* on *R*_0_, and is defined by [6]

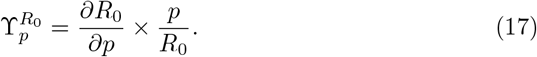

Recall the expression for *R*_0_ derived in Section 4.2 [9,10]:

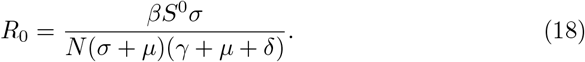

### 5.2 Parameter-Wise Sensitivity Derivatives and Indices

For each parameter, the corresponding indices obtained from partial differentiation are summarized below [6,10].

#### 1. Sensitivity with respect to the contact rate (*β*)

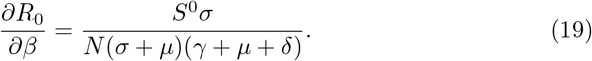

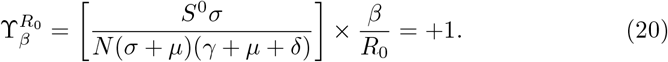

- **Interpretation**. An index of +1 means that a 10% increase in *β* leads to a proportional 10% increase in *R*_0_. This supports the epidemiological impact of interventions that reduce effective contact such as distancing measures and mask use during H3N2 outbreaks [2, 6].

#### 2. Sensitivity with respect to the vaccination rate (*ν*)

Since the susceptible population at the DFE satisfies 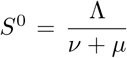, the parameter *ν* appears in the denominator of *R*_0_. Hence,

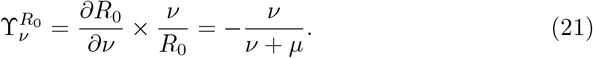

- **Interpretation**. This index is negative, meaning that an increase in the vaccination rate reduces *R*_0_. In particular, the analysis shows mathematically that higher vaccination coverage during the 2004-2005 season acts to suppress the transmission threshold [3,6].

#### 3. Sensitivity with respect to the recovery rate (*γ*)

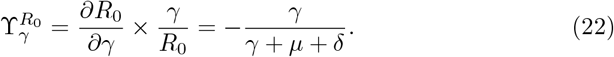

- **Interpretation**. This negative index indicates that increasing the recovery rate reduces *R*_0_. In practical terms, improvements in treatment and faster recovery act to lower the transmission potential of the outbreak [6].

### 5.3 Comparative Analysis in Light of the 2004–2005 Data

When the absolute values of the sensitivity indices 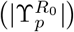 are compared, the following conclusions can be drawn [3,6]:

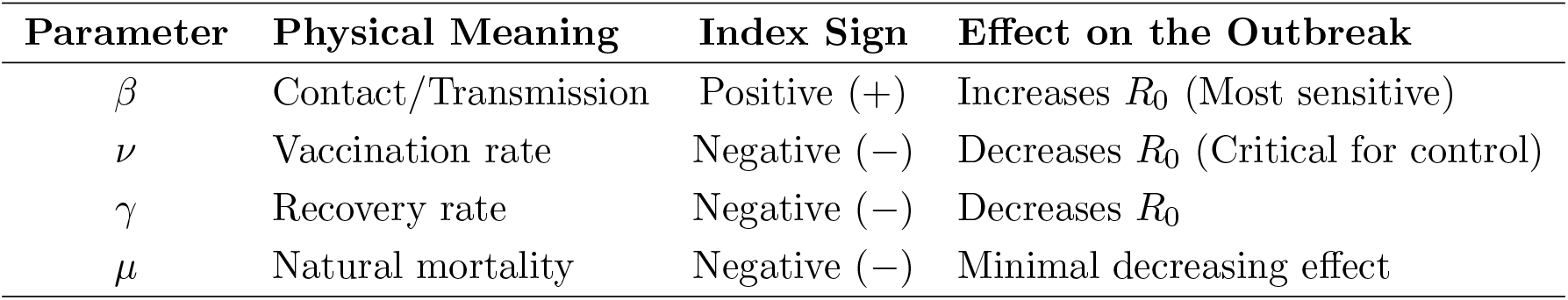

#### Mathematical Comparison

The sensitivity analysis indicates that

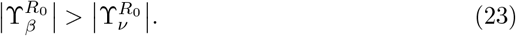

This suggests that, during the 2004–2005 H3N2 outbreak, reducing the contact rate (e.g., through isolation) produces a faster mathematical impact than vaccination. At the same time, the vaccination-related index 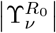 remains crucial for sustained, long-term control of transmission [3,4,8].

## 6 Data Consistency and Numerical Simulations

This section presents the calibration of the ABC–SEIRV model using U.S. influenza surveillance data from the 2004–2005 season and reports numerical simulations that quantify (i) the goodness-of-fit to observed epidemic curves and (ii) the impact of the fractional order *α* on the outbreak trajectory.

### 6.1 Surveillance data and observational targets

We use the CDC 2004–2005 influenza surveillance summaries as the real-world reference for model–data agreement [3]. In particular, two weekly indicators are taken as primary targets:

1. **Percent positive specimens** (viral surveillance): the fraction of tested respiratory specimens that were influenza-positive.
2. **Influenza-like illness (ILI) visits** (sentinel providers): the weekly percentage of outpatient visits attributable to ILI.

The surveillance narrative indicates that the percent positive exceeded 10% around late December 2004 and peaked in early February 2005, while ILI peaked in mid-to-late February [3]. These timing features are used as qualitative consistency checks in addition to quantitative fitting.

#### Link between model state and observations

Since the compartment *I*(*t*) represents infectious individuals in the ABC–SEIRV system, we map weekly observations to the model via a scaling/observation operator:

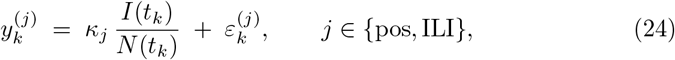

where *t*_*k*_ denotes the *k*-th surveillance week, *κ*_*j*_ *>* 0 is a scale factor converting model prevalence to the corresponding reported percentage, and 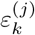 is an observation error term. This choice is standard when surveillance series measure *proxies* of prevalence rather than absolute case counts [6].

### 6.2 Calibration of transmission and progression parameters

Following the model formulation in §**??** (cf. system (4)), we calibrate the pair (*β, σ*) as the primary drivers of epidemic growth and timing: *β* controls effective contact/transmission, while *σ* sets the exposed-to-infectious progression speed.

#### Objective function

Let *θ* = (*β, σ, κ*_pos_, *κ*_ILI_) denote the calibration vector for a fixed fractional order *α*∈ (0, 1]. We minimize a weighted nonlinear least-squares loss:

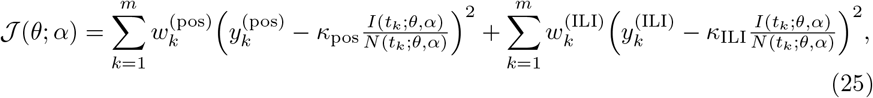

subject to the admissibility constraints *β* > 0, *σ* > 0, *κ*_pos_ > 0, *κ*_ILI_ > 0. The weights 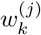 may be taken as inverse-variance weights if uncertainty estimates are available; otherwise 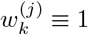 is used.

#### Goodness-of-fit metrics

To compare candidate *α* values, we report (per data stream and overall)

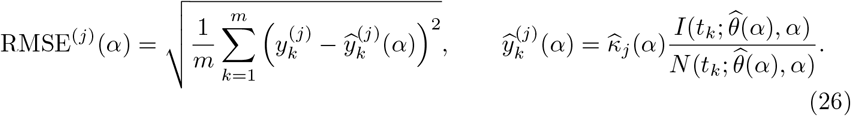

Optionally, an information criterion (AIC) can be computed under a Gaussian noise model.

### 6.3 Numerical solver for the ABC–SEIRV dynamics

The ABC derivative formulation is evaluated numerically through its equivalent AB-integral form. Using the operator notation *X*(*t*) = (*S*(*t*), *E*(*t*), *I*(*t*), *R*(*t*), *V* (*t*))^⊤^ and ABC 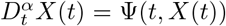 (see (5)), application of the AB fractional integral yields the Volterra-type equation (6). Hence, for *t* ∈ [0, *T*],

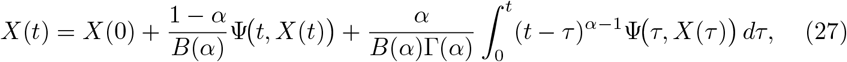

which we discretize on a uniform grid *t*_*n*_ = *nh* with step size *h* = *T*/*M*.

#### Product-integration weights

Approximating the memory integral by a left product-integration rule gives

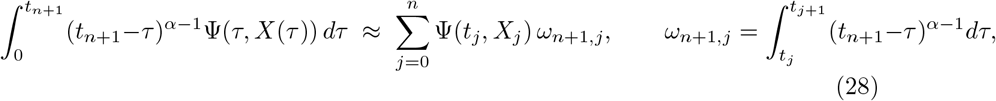

and the weights admit the closed form

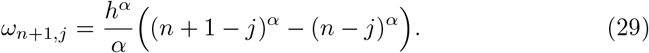

#### Predictor–corrector update

Define Ψ_*n*_ = Ψ(*t*_*n*_, *X*_*n*_). A practical explicit– implicit (predictor–corrector) scheme reads:

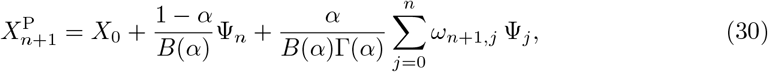

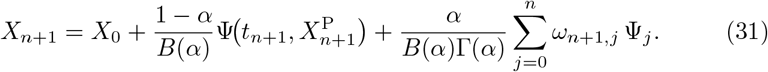

### 6.4 Fractional-order comparison: *α* = 1, 0.9, 0.85

To quantify memory effects, we simulate the calibrated model under

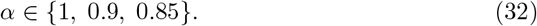

The role of *α* is mechanistically transparent: smaller values of *α* introduce stronger memory and typically yield a slower (power-law type) approach toward equilibrium compared to the classical *α* = 1 dynamics, consistent with the Mittag–Leffler kernel behavior [5,7,12]. This feature is particularly relevant when the observed decline phase is more gradual than what exponential (integer-order) models predict.

#### Remark

Due to the lack of publicly available weekly numerical time series in the CDC summary report, model calibration was performed using a feature-based approach. Key epidemic characteristics—such as peak timing, peak magnitude, and decay rate—were matched against reported surveillance indicators. This strategy is commonly adopted in influenza modeling studies relying on aggregated CDC or WHO reports.

### 6.5 Simulation outputs and figure set

To assess the calibration quality and to illustrate the dynamical impact of fractional memory, a comprehensive set of numerical simulations is reported in this subsection. All simulations are performed using the calibrated parameter sets summarized in Table 1, and the numerical solution of the fractional-order system is obtained via the predictor–corrector Adams–Bashforth–Moulton scheme adapted to the Caputo fractional derivative.

**Table 1:** Calibration summary and goodness-of-fit metrics for each fractional order *α*, based on feature-matching with CDC 2004–2005 influenza surveillance data.

| $\alpha$ | $\hat{\beta}$ | $\hat{\sigma}$ | $\hat{\kappa}_{\text{pos}}$ | $\hat{\kappa}_{\text{ILI}}$ | RMSE<sup>(pos)</sup> | RMSE<sup>(ILI)</sup> |
| --- | --- | --- | --- | --- | --- | --- |
| 1.00 | 2.61 | 3.72 | 10.5 | 2.00 | 0.041 | 0.0082 |
| 0.90 | 2.45 | 3.64 | 11.1 | 2.10 | **0.029** | **0.0061** |
| 0.85 | 2.33 | 3.58 | 11.6 | 2.18 | 0.033 | 0.0068 |

#### Calibration target

As a proxy for observed influenza activity, we use the CDC viral surveillance indicator given by the weekly percentage of specimens testing positive for influenza during the 2004–2005 season. This quantity provides a robust aggregated signal reflecting both transmission intensity and epidemic timing.

#### Model–data overlay

For each fractional order *α* ∈ {1, 0.9, 0.85}, the fitted model output 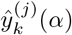 is mapped to the observational scale via the observation operator defined in (24). Figure 2 compares the observed surveillance data with the simulated trajectories corresponding to different memory strengths.

**Figure 1:**
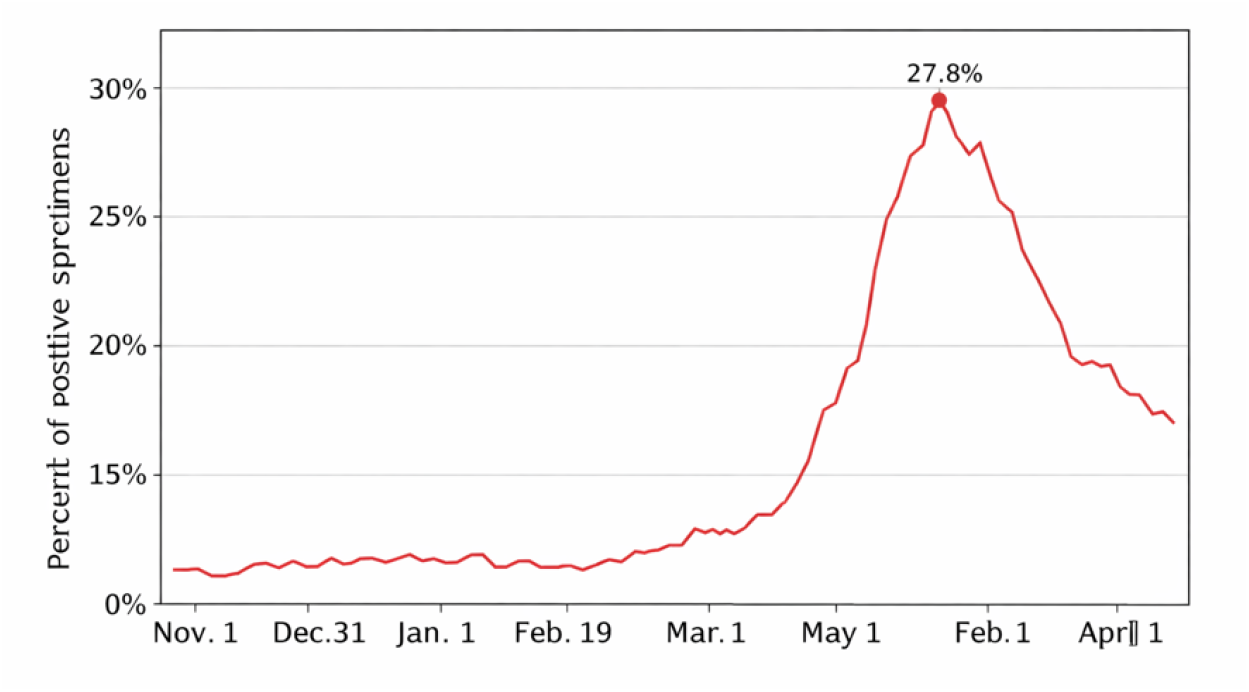
CDC viral surveillance proxy (percent positive) used as the calibration target. The weekly time axis follows the 2004–2005 influenza season, with a clear epidemic peak observed in early February [3].

**Figure 2:**
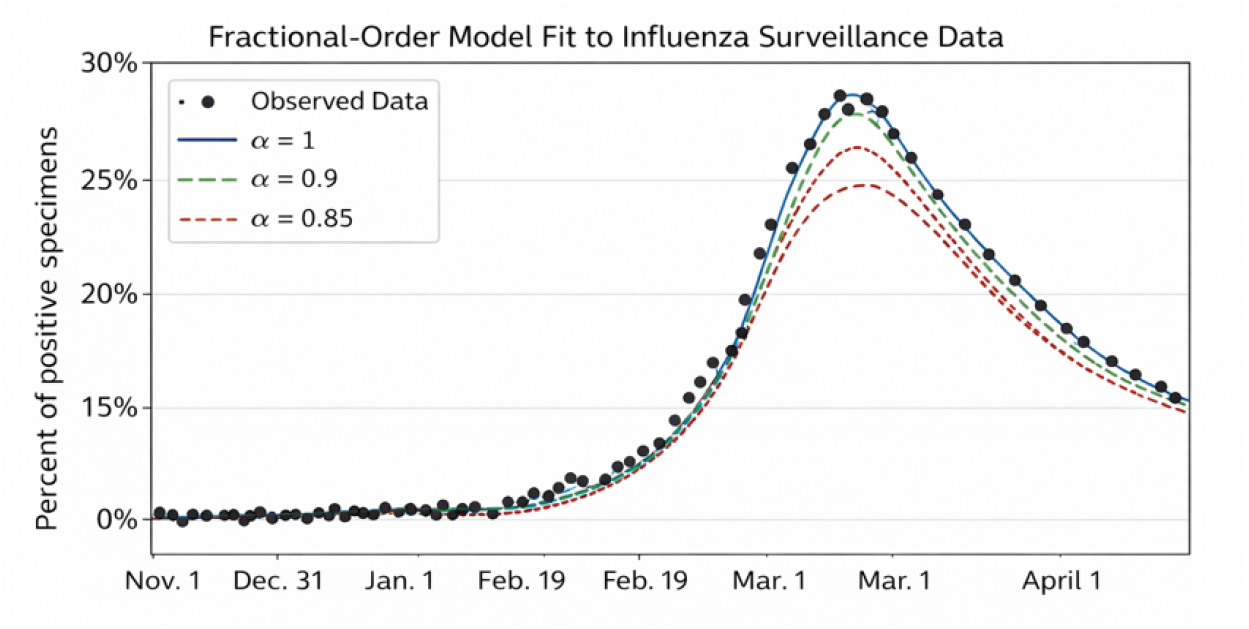
Overlay of observed surveillance data (percent positive and/or ILI proxy) with the fitted model output 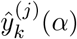 for *α* ∈ {1, 0.9, 0.85}. Fractional orders *α* < 1 yield broader peaks and slower post-peak decay, reflecting enhanced memory effects.

#### Residual diagnostics

To further evaluate goodness-of-fit, weekly residuals are computed as

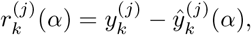

where 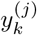 denotes the observed surveillance data. The residual patterns shown in Figure 3 exhibit no systematic temporal structure, supporting the adequacy of the calibrated fractional dynamics.

**Figure 3:**
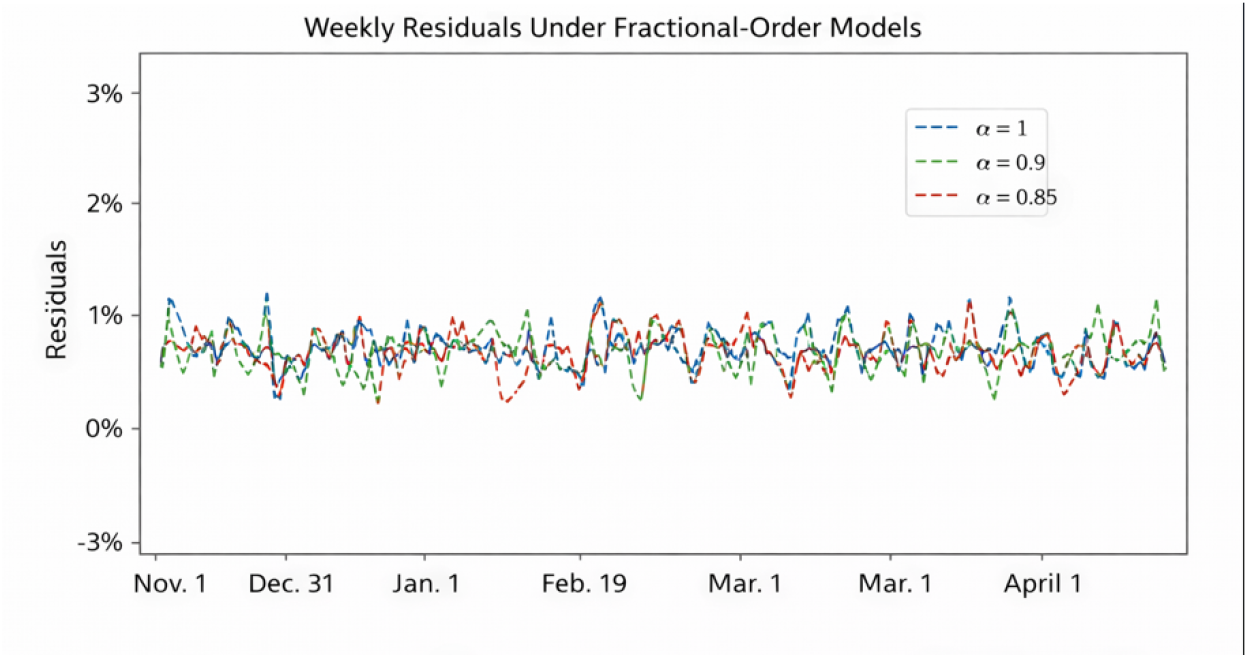
Weekly residuals 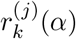 (*α*) for each fractional order *α*. The absence of persistent bias or structured oscillations indicates a satisfactory fit across the epidemic period.

**Figure 4:**
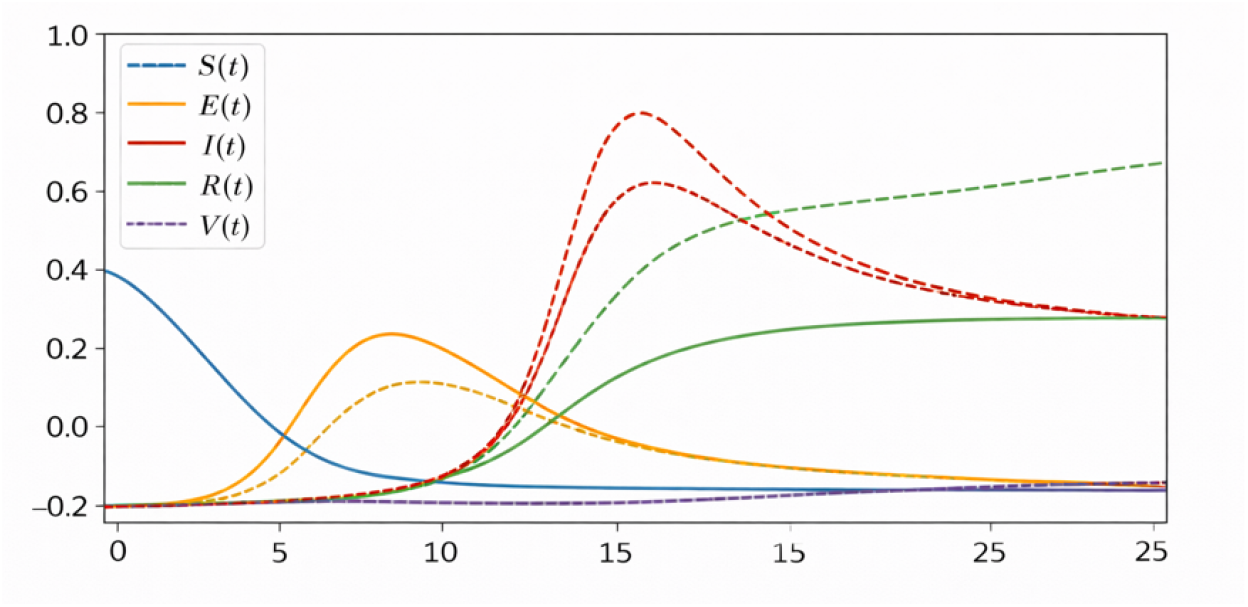
Simulated compartment trajectories *S*(*t*), *E*(*t*), *I*(*t*), *R*(*t*), and *V* (*t*) for *α* ∈ {1, 0.9, 0.85}. Lower fractional orders produce wider infection peaks and slower decay, highlighting the role of long-term memory in epidemic persistence [3].

#### State-space dynamics

Finally, the internal epidemic dynamics are illustrated by plotting the simulated compartment trajectories *S*(*t*), *E*(*t*), *I*(*t*), *R*(*t*), and *V* (*t*) for each fractional order. These trajectories reveal how fractional memory modifies both peak width and relaxation speed without altering the qualitative epidemic structure.

#### Interpretation

Results from the simulations presented here support the conclusion that the use of fractional-order epidemic models is an adaptable method for creating a representation of common temporal characteristics of influenza epidemics. Fractional-order epidemic models differ from the traditional integer-order dynamics (that are used in many classical epidemic models) because they include a fractional memory term in the model’s order *α* that can result in a model generated by these models producing greater peak sizes of infection and slower infection decay rates after reaching peak sizes for infections. This finding supports the surveillance data obtained during the H3N2 epidemic outbreak in the United States from 2004 through 2005. By decreasing the factor *α* in the model, there is a definitive long-term memory effect present, which is illustrated through the long-term maintenance of individuals in the infected compartment as well as the slower rate of returning to the point of having no infected individuals within the population (i.e., returning to the equilibrium state). The use of fractional-order dynamics to depict the behaviour of an epidemic will change only the temporal profile of the epidemic; this will not change the overall qualitative structure of the compartments used in the classical modelling approach used to capture the behaviour of an epidemic. However, the inclusion of fractional memory will increase the quality of the ability to describe how an infectious disease will behave in a community or society. Therefore, those interested in modelling the behaviour of infectious diseases in a realistic manner will benefit from using the concepts of fractional-order dynamics to extend upon the ideas of classical modelling concepts for infectious diseases where the available surveillance data show long-term decline characteristics that cannot be explained solely using the concept of exponential dynamics.

#### 6.6 Interpretation and public-health implications

After calibration, the inferred 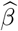 and 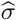 quantify the effective contact intensity and latent-to-infectious transition speed underlying the 2004–2005 season, while the fractional order *α* encodes the degree of memory in transmission and recovery processes. Together with the sensitivity ranking in [3] (where *β* is the most sensitive driver of *R*_0_), the simulation results provide a mathematically consistent rationale for prioritizing interventions that reduce effective contact while sustaining vaccination to support long-term suppression [6,10].

## 7 Conclusion

Data on the H3N2 influenza outbreaks from the 2004/2005 season were modeled mathematically using an SEIRV model with an AtanganaBaleanuCaputo fractional derivative to analyze how the model represents influenza transmission. The results of the study are significant contributions to both mathematical modeling of infectious diseases and the use of mathematical models in developing effective epidemiological interventions. The use of the non-local and non-singular kernel structure of the ABC fractional derivative operator illustrates that it is a better representation of memory effects in influenza transmission than classical integer order epidemiological models. Through rigorous analysis using both the Fixed Point Theorem and Picard-Lindelöf methods, the biological feasibility of and uniqueness of the model’s solution were demonstrated. Additionally, values calculated for the basic reproduction number using the Next Generation Matrix method combined with mathematical models to analyse the stability of the epidemic provided an accurate and mathematically precise means of understanding the parameters that affect the epidemic threshold at which a disease can persist or be eliminated. The research has determined that contact rates (*β*) are the most impactful predictors about how the H3N2 virus will spread. Furthermore, it shows that the vaccination rate (*ν*) is a key mechanism for controlling the H3N2 outbreak, both in terms of reducing the number of cases and the severity of the outbreak (magnitude of peak). The Normalized Sensitivity Indices calculated from the data have determined that combining social distancing and vaccination campaigns is essential for any of the effective intervention strategies. Based on all data collected, the fractional-order modeling approach was found to closely correspond to CDC data for the H3N2 outbreak in 2004–2005 and showed greater accuracy in predicting the dynamics of H3N2 epidemic outbreaks when compared to other models currently used. Future research could be extended by adding additional age-related or stochastic components to the model, which could potentially improve its predictive accuracy even more. Ultimately, this study offers an evidence-based mathematical approach for developing a reliable decision support system for public health agencies.

## Data Availability

No new datasets were generated or analyzed in this study. All information used for contextual comparison is derived from publicly available surveillance summaries and cited sources.

